# Implementing the mhGAP-HIG: The process and evaluation of training primary health care workers in Khyber Pakhtunkhwa, Pakistan

**DOI:** 10.1101/2024.11.21.24317704

**Authors:** Asma Humayun, Arooj Najmussaqib

## Abstract

**Background:** Building the capacity of primary care staff is crucial for integrating mental health care into primary healthcare to reduce the significant treatment gap for psychiatric disorders prevalent in low- and middle-income countries. This research investigates the effectiveness of adapted mhGAP-HIG guidelines in Khyber Pakhtunkhwa (KP).

**Methods:** A team of seven trained experts conducted six five-day training workshops in six districts of KP. A total of 105 primary care physicians and clinical psychologists were trained. Mix method analysis was performed. Paired sample t-tests were used to compare knowledge scores pre- and post-training and after 8 months. Thematic analysis was conducted to examine feedback of the participants regarding the training, whereas content analysis was performed on the reflections of the trainers on the adapted guide.

**Results:** Findings demonstrated significant improvements in knowledge of the participants to identify and manage common mental health conditions. The percentage of correctly answered questions in the pre-test was 73.86%, which increased to 85.94% in the post-test, indicating a 12.08% increase in knowledge. Most noticeable improvements in knowledge were observed in harmful use of substances (22.56), General principles of care and other significant mental health complaints (15.15%), stress (13.80%), suicide, and epilepsy (13.13%). The thematic analyses highlighted the strengths and gaps of training and made recommendations to strengthen preservice training and provide regular refresher courses.

**Conclusion:** The study underscores the feasibility of implementing an adapted mhGAP-HIG for training primary care physicians and clinical psychologists within existing healthcare resources of KP. The preliminary findings endorse the scalability across other provinces in the country.

## Introduction

Pakistan has a devolved federal structure, where health is a provincial subject. Each province has struggled to develop mental health either because of their ongoing need to respond to other competing public health challenges and a dearth of mental health expertise and resources (1).

The province of Khyber Pakhtunkhwa (KP), located in the north-western region of Pakistan, is divided into 37 districts. Over the last three decades, the province has borne the brunt of conflict and terrorism, natural disasters, internal displacements, and hosting a refugee population of 1.8 million. It is estimated that only 30% of KP’s women and children have access to medical services due to cultural barriers and other socio-economic restrictions (2) Additionally, worrying suicide rates have been reported in some districts like Chitral, which lacks any psychiatric services (3,4).

In 2023, the Ministry of Planning Development & Special Initiative (MoPD&SI) provided technical support to the Directorate of Public Health in KP to conduct a pilot project for building the capacity of primary health care workers (PHCWs), which included primary care physicians (PCPs) and clinical psychologists (CPs). The project was supported by the International Medical Corps (IMC) to strengthen mental healthcare services for Afghan refugees and host communities in nine districts (Chitral, Haripur, Kohat, Lower Dir, Mansehra, Mardan, Nowshera, Peshawar, and Swabi).

## Background context

The MoPD&SI developed an evidence-based and scalable model for a multi-layered mental healthcare system in 2021 (5). This model aims to strengthen the existing mental healthcare system by utilizing evidence-based resources to build the capacity of a mental health workforce outside tertiary care hospitals; supervise this workforce to provide mental health and psychosocial support services in primary care; establish a referral pathway and collect relevant data. As part of the digital tools for this model, the mhGAP Humanitarian Intervention Guidelines (mhGAP-HIG) (6) were adapted and developed into a mobile application (mhGAP-HIG-PK tools) for Pakistan (7).

In contrast to earlier mhGAP training initiatives, which either implemented the guide without modifications (8,9) or partially adapted it (10–12), the mhGAP-HIG-PK has been comprehensively adapted and contextualized for Pakistan. Furthermore, its clinical utility has been enhanced by integrating evidence-driven interview questions for assessment protocols and step-by-step techniques for delivering psychosocial interventions. The mobile application offers a complete reference to mhGAP-HIG-PK and facilitates the submission of clinical data by trained PHCWs, along with the provision to seek supervision when needed.

Similar to other LMICs, major challenges for implementing the mhGAP guidelines in Pakistan include a dearth of specialist resources, disease-centered practices, inadequate referral pathways, knowledge and skills gaps pertaining to pre-service training of PCP, and stigma towards people with mental disorders (7,13).

This paper discusses the training process and evaluation using the mhGAP-HIG-PK tools. A subsequent paper will discuss post-training supervision and the analysis of the clinical data collected during supervision.

## Methods

This study used a mixed-methods approach, integrating quantitative and qualitative data collection and analysis. This study was conducted as part of the Mental Health and Psychosocial Support Project,approved by Ministry of Planning, Development and Special Initiatives in compliance with ethical standards and consent protocols under letter no. 6(262) HPC/2020.

### 1. Analysis of mental health services in KP

This phase aimed to determine the current condition of mental health services, with a particular emphasis on understanding the challenges encountered by PHCWs while dealing with mental health issues. To accomplish this, we conducted two Focus Group Discussions (FGDs) and examined existing literature to gain a comprehensive understanding of the dynamics of mental health care in KP. The FGDs had 15 participants, including PCPs, CPs, IMC staff, and KP health directorate representatives. The purpose of the FGDs was to gather the perspectives of PHCWs on their experiences and the difficulties presented by local culture and regulatory frameworks. The insights reported here are the result of an in-depth review of the information gathered through these interactions.

There exists a critical shortage and uneven distribution of mental health services in KP. For a population of 40 million, there are approximately 50 psychiatrists (1 psychiatrist per 800,000 individuals), with nearly half located in Peshawar, the provincial capital. Only the urban clusters (nine districts) have any psychiatric services, whereas over 81% of the population lives in rural areas (14). The gap in mental healthcare in the province is not only limited to a dearth of services, but the quality of care is also a major concern. Psychiatric services are mainly bio-medical in approach.

The CPs are an important resource in the province, but there are only a few positions in tertiary care, public hospitals, and academic institutions. Over a hundred CPs are working in other settings, such as humanitarian (non-government) organizations, rehabilitation centers, or the private sector. Unfortunately, there is a severe dearth of opportunities either for seeking supervision or to strengthen their skills as most of them work in project-based, short-term positions (15).

The Health Information System in KP includes three mental disorders (Depression, Epilepsy, and Drug dependence). Despite existing mechanisms to report data, no information is ever reported. Furthermore, psychiatry education within the MBBS curriculum encounters several challenges, such as minimal implementation despite its mandatory status, the absence of a dedicated examination, a shortage of qualified faculty, and inadequate pre-service training, all of which contribute to reduced student motivation (16).

### 2. Selection and training of trainers

Selecting trainers posed challenges as most of the qualified and experienced mental health professionals had full-time academic positions and private practice. Through our professional networks, we identified potential trainers from or close to our target districts. The selection criteria also encompassed attributes such as integrity, motivation for community service, and proficiency in a biopsychosocial approach, extending beyond the traditional biomedical model. The selected trainers volunteered to participate in the training activity, as no financial incentive or official leave from work was provided.

A team of eight trainers (5 psychiatrists and 3 CPs) was selected and trained. One of them dropped out because of logistic concerns. Due to the trainers’ diverse geographic locations spanning numerous districts and limited resources, it was not feasible to arrange an in-person training session. Consequently, the training was conducted online over three weeks under the supervision of a master trainer. Trainers were given a digital copy of the mhGAP-HIG-PK guide during the first week to familiarize themselves with the detailed modules. During the following week, they were required to focus on preparing just one module. In the third week, every participant facilitated a one-hour teaching session. Two external reviewers were invited to provide objective feedback on both the content of the presentations and the instructional methods used to improve the evaluation process.

The second part of their training involved practical supervision during the training workshops. Every training workshop was led by a minimum of 2 trainers. To facilitate the learning process and foster interactive peer feedback, a WhatsApp group was formed. This platform was utilized for the daily exchange of updates, recorded videos of role-plays, and feedback from peers and trainees. Upon completion of training workshops, trainers were awarded digital certificates to recognize their contribution.

### 3. Selection and training of PHCWs

The Directorate of Public Health engaged with the relevant district health authorities to nominate PCPs for the training program. Similarly, IMC nominated CPs from within their teams or those affiliated with partner agencies for participation. No recruitment criteria were specified for PCP since they were nominated by the health department, but a criterion based on level of formal education was developed for CPs (either a MS degree or advanced diploma in clinical psychology).

### 4. Training workshops

Due to logistic constraints, instead of nine districts, these were merged to hold a total of six 5-day training workshops at Chitral; Haripur (for districts of Haripur and Mansehra); Lower Dir, Mardan 1 (Mardan and Swabi); Mardan 2 (Mardan and Nowshera); and Peshawar (Peshawar and Kohat) between August-December 2023. Following each training workshop, the trainees were supervised for three months.

To accommodate participants’ lengthy commutes, daily sessions were reduced to 5.5 hours, with an added half-hour for home study. This structure resulted in a cumulative training duration of 30 hours over five days. All attendees were required to register on the Mental Health and Psychosocial Support (MHPSS) web portal, including the Learning Management System (LMS). The training sessions were conducted using interactive learning methodologies, incorporating role-plays, small group discussions, and reflection activities to enhance engagement and comprehension. The training resources included mhGAP-HIG-PK tools (hard copy and a mobile application).

Each day started with a 30-minute ‘recap session’ of the prior day’s modules. The programme covered all eleven mhGAP-HIG modules, including General principles of care (GPC), Acute stress (ACU), Grief (GRI), Depression (DEP), PTSD, Psychosis (PSY), Epilepsy (EPI), Intellectual disability (ID), Harmful use of alcohol and drugs (SUB), Suicide (SUI), and Other Significant Mental Health Complaints (OTH). The average duration for each module was 90 minutes. Upon successful completion of the training, trainees were awarded digital certificates to recognize their achievement.

### 5. Refresher Trainings

Due to logistical and budgetary constraints, only three refresher training workshops were conducted covering six districts—Chitral, Kohat, Mardan, Nowshera, Peshawar, and Swabi - eight months following the initial training. A total of 48 participants attended these refresher trainings. The areas identified with participants with an aim to strengthen their skills included assessment of depression, suicide risk, and children with intellectual disability. Additionally, psychosocial interventions were practiced in role plays for common conditions, including acute stress (especially bed wetting, dissociation), grief, depression, intellectual disability, epilepsy, and others. Furthermore, discussions on the pharmacological interventions for depression, psychosis and epilepsy were addressed knowledge gaps.

### 6. Pre and post training assessment

The pre- and post-test assessments were conducted using an adapted knowledge questionnaire (7) comprising 25 items. For analysis, these items were categorized into clusters corresponding to mhGAP-HIG modules: GPC (2 items), ACU (3), Grief (2), PTSD (1), DEP (3), PSY (3), ID (2), SUB (3), SUI (2), EPI (2), and Others (2). The assessment was repeated after 8 months to measure knowledge retention.

### 7. Feedback by the trainees

PHCWs submitted quantitative and qualitative feedback through an online questionnaire on the Learning Management System (LMS) upon completing the training.

### 8. Reflections by the participants on the process of training

Collective reflections by the participants (trainees and trainers) were summarized in written reports by the respective trainers at the end of each day. These reports drew on their observations and discussions from daily ‘recap sessions’ with trainees.

## Data Analysis

The data was cleaned and coded before conducting analysis using SPSS version 26 (17). The descriptive statistics were reported as percentages or as the mean with its corresponding standard deviation. 105 participants attended the training; however, six questionnaires were discarded due to missing information. For analysis purposes, data from 99 participants was organized by district. We also conducted knowledge comparisons between the pre- and post-assessment using clusters. Given the normal distribution of the data, a paired t-test was performed using the ‘pre-,’ ‘post-’, and ‘8 months after’ mean scores to evaluate any changes in the knowledge scores following the training. Furthermore, qualitative feedback from the training participants and trainers’ reflections were analyzed using thematic and content analysis respectively.

## Results

This section presents the demographics, pre and post training analyses, feedback and reflections of the participants. The trainees comprised 64% men with nearly half having at least 5 years of work experience and 64% working in primary care settings, indicating varied qualification, diverse expertise and reflections brought to this training. The demographic details of all participants are shared in Table 1.

**Table 1:** Demographics of trainees (N=105)

|  | **Doctors** | **Psychologists** |
| --- | --- | --- |
| **Gender** | **n (%)** | **n (%)** |
| Males | 49 (47%) | 18 (17%) |
| Females | 25 (24%) | 13 (12%) |
| **Experience** |  |  |
| <5 years | 17 (16%) | 14 (13%) |
| 5-10 years | 37 (35%) | 12 (11%) |
| >10 years | 20 (19%) | 5 (5%) |
| **Healthcare level** |  |  |
| Primary (iNGO) | 38 (36%) | 29 (28%) |
| Secondary | 35 (33%) | 2 (2%) |
| Tertiary | 1 (1%) |  |
| **Education level (Doctors)** |  |  |
| MBBS | 66 (89%) |  |
| MBBS + MPH | 3 (4%) |  |
| MBBS+MCPS (Medicine, ENT) | 3 (4%) |  |
| MBBS+FCPS/MD (Internal Medicine) | 2 (2%) |  |
| **Education level (Psychologists)** |  |  |
| MS Clinical Psychology | 13 (42%) |  |
| Advanced Diploma in Clinical Psychology | 15 (48%) |  |
| BS Clinical Psychology | 3 (10%) |  |

### 1. Pre- and post-training outcomes

The analysis of pre- and post-training outcomes suggested encouraging levels of baseline knowledge. The percentage of correctly answered questions in the pre-test was 73.86%, which increased to 85.94% in the post-test, indicating a 12.08% increase in knowledge.

Table 2 demonstrates meaningful improvements in knowledge levels across several regions following training. There was a significant increase in mean knowledge scores between the pre- and post-training for every district. However, Mardan1 shows an increase in knowledge score post-training, though with a marginally higher significance level. This implies a positive but less definitive impact of the training. Furthermore, Peshawar exhibited the most pronounced enhancement (p=<.001).

**Table 2:** Pre and Post Knowledge Comparisons by Districts (N=99)

| Area | Pre<br>M | SD | Post<br>M | SD | Paired Test |  | After 8 months |  |  |
| --- | --- | --- | --- | --- | --- | --- | --- | --- | --- |
|  |  |  |  |  | t value | P | M | SD | P |
| Haripur | 19.63 | 2.53 | 22.50 | 1.79 | 3.439 | 0.004 |  |  |  |
| Chitral | 18.07 | 3.95 | 21.29 | 3.77 | 2.527 | 0.025 | 20.3 | 2.75 | 2.37 (.042) |
| Lower Dir | 17.69 | 3.34 | 21.25 | 2.65 | 3.578 | 0.003 | - | - | - |
| Mardan1 | 19.30 | 2.83 | 21.10 | 2.53 | 1.911 | 0.071 | - |  |  |
| Mardan2 | 17.88 | 3.20 | 21.50 | 1.51 | 4.389 | 0.001 | 21.42 | 2.68 | 3.82 (.002) |
| Peshawar | 18.00 | 2.32 | 21.35 | 2.29 | 4.968 | <0.001 | 20.90 | 2.70 | 4.03 (.001) |

The decrease in standard deviations across districts post-intervention indicates a reduction in variability in participants’ knowledge scores. This implies that disparities in knowledge have narrowed, leading to the attainment of a more uniform level of understanding or expertise among participants after the training, as has been previously reported (18).

Furthermore, in the districts of Chitral, Peshawar and Mardan2, mean knowledge scores significantly improved, reaching 20.3 (SD=2.75), 20.90 (SD=2.70), and 21.42 (SD=2.68) respectively [pre-training versus 8 months post-training (Table 2)]. This highlights the training’s effectiveness in retention of knowledge over time, consistent with literature (11,19).

Table 3 demonstrates the improvement in participants’ knowledge comparisons by clusters. Overall, knowledge of the participants significantly improved from 18.46 to 21.48 (p<.001). Furthermore, each cluster also showed significant improvement in all modules except Grief and ID.

**Table 3:** Knowledge Comparisons by Clusters (N=99)

|  | Pre |  | Post |  | Paired t-Test |  |
| --- | --- | --- | --- | --- | --- | --- |
| Cluster | M | SD | M | SD | t value | P |
| GPC | 1.10 | 0.65 | 1.40 | 0.60 | 3.545 | 0.001 |
| Stress | 2.12 | 0.75 | 2.54 | 0.61 | 4.252 | <.001 |
| Grief | 1.90 | 0.30 | 1.95 | 0.22 | 1.393 | 0.167 |
| PTSD | 0.80 | 0.40 | 0.89 | 0.32 | 26.21 | <.001 |
| Depression | 2.28 | 0.78 | 2.53 | 0.61 | 2.362 | 0.02 |
| Psychosis | 2.46 | 0.61 | 2.74 | 0.53 | 3.089 | 0.003 |
| ID | 1.51 | 0.61 | 1.65 | 0.50 | 1.861 | 0.066 |
| SUD | 2.09 | 0.72 | 2.77 | 0.49 | 7.767 | <.001 |
| Epilepsy | 1.36 | 0.68 | 1.63 | 0.51 | 3.483 | 0.001 |
| Suicide | 1.28 | 0.54 | 1.55 | 0.56 | 3.766 | <.001 |
| Others | 1.56 | 0.67 | 1.86 | 0.38 | 4.268 | <.001 |

The module with the most noticeable improvements in scores was SUB (22.56%). This is followed by GPC and Others (15.15%), Acute stress (13.80%), Suicide and Epilepsy (13.13%), PTSD, and Psychosis (9.09%). On the other hand, the ID and Grief modules observed insignificant improvements, with percentage increases of 7.07% and 2.53%, respectively.

### 2. Feedback

Trainee’s feedback was analyzed through rating forms and qualitative responses. Their feedback provides insights into the program’s overall success and areas for enhancement (see Supplementary file). Notably, 75% rated their experience as excellent and 23% as good, indicating that the program was highly effective and well-received.

#### Qualitative feedback

The thematic analysis from the feedback indicates that the participants were highly satisfied with the educational experience and appreciated the content as well as the interactive pedagogy. The training appeared effective in strengthening knowledge and clinical skills, particularly to identify and manage mental health conditions.

Overall, the trainees expressed satisfaction with the training, deeming it both beneficial and informative. They particularly valued the interactive methodologies used, such as role-plays and engaging group discussions. One participant highlighted the effectiveness of these methods, stating, “The interactive method of training made the learning multifold.” In addition to the instructional approach, participants were appreciative of the provision of hard copies of the guide, which enhanced their learning experience.

Concerning the impact on knowledge and clinical skills, the participants reported significant enhancements in their understanding and abilities related to the identification, diagnosis, and treatment of mental disorders, incorporating holistic, psychosocial, and pharmacological approaches. The training also improved their capabilities in making management plans, with one participant noting, “It became clearer whom to refer and whom to treat, who needs no treatment, who needs counseling, and who needs pharmacological intervention.”

The trainees recognized and appreciated the expertise of senior trainers. They advocated for the broader dissemination of mhGAP trainings for all healthcare and community workers. They also recommended organizing refresher courses to refine their skills. Furthermore, the integration of the mhGAP guide into the medical curriculum was suggested.

### 3. Reflections by the trainers and feedback by the trainees

The outcome of the analysis of the reflections by the trainers and feedback by the trainees on individual modules is presented under three categories in Table 4:

**Table 4:**
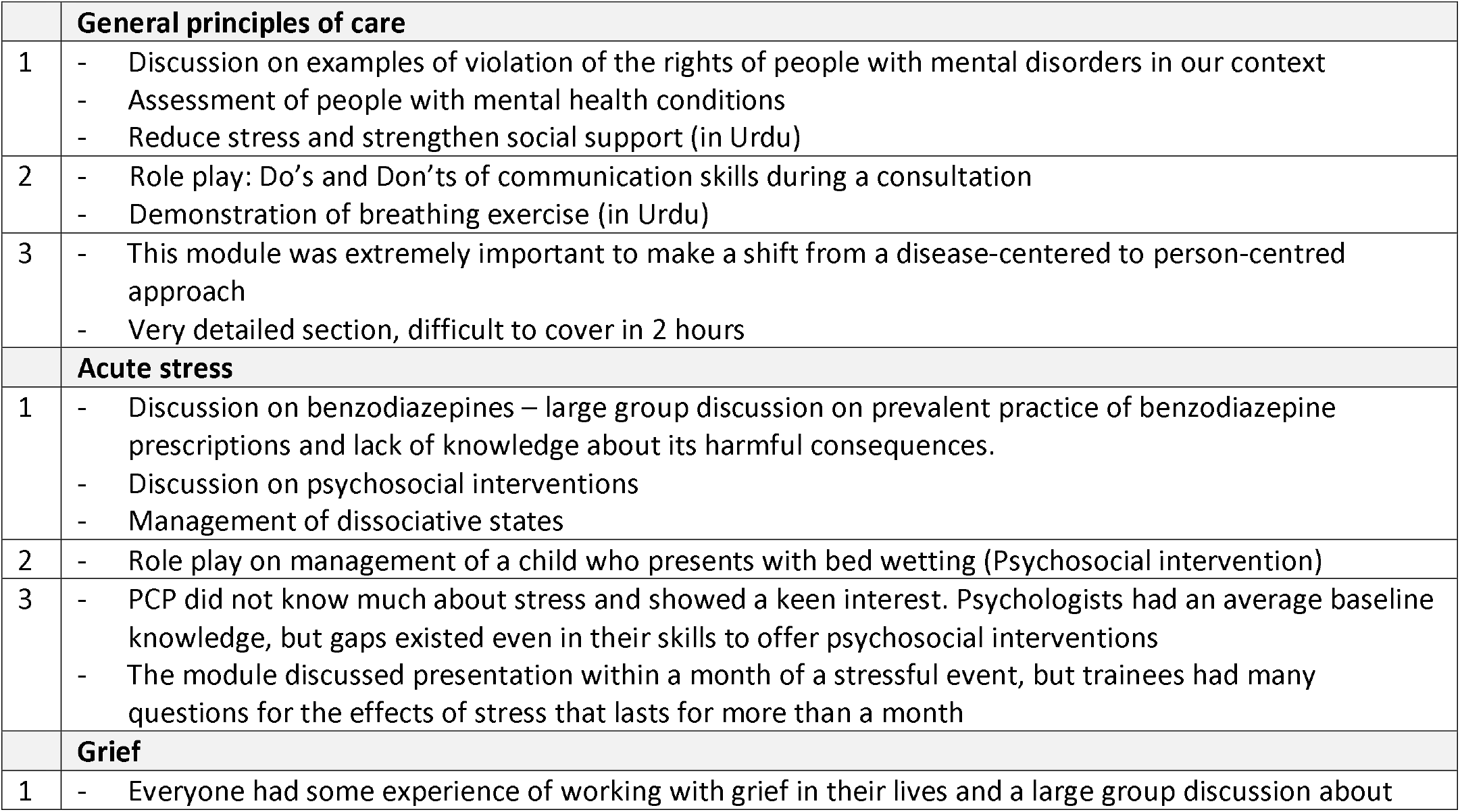
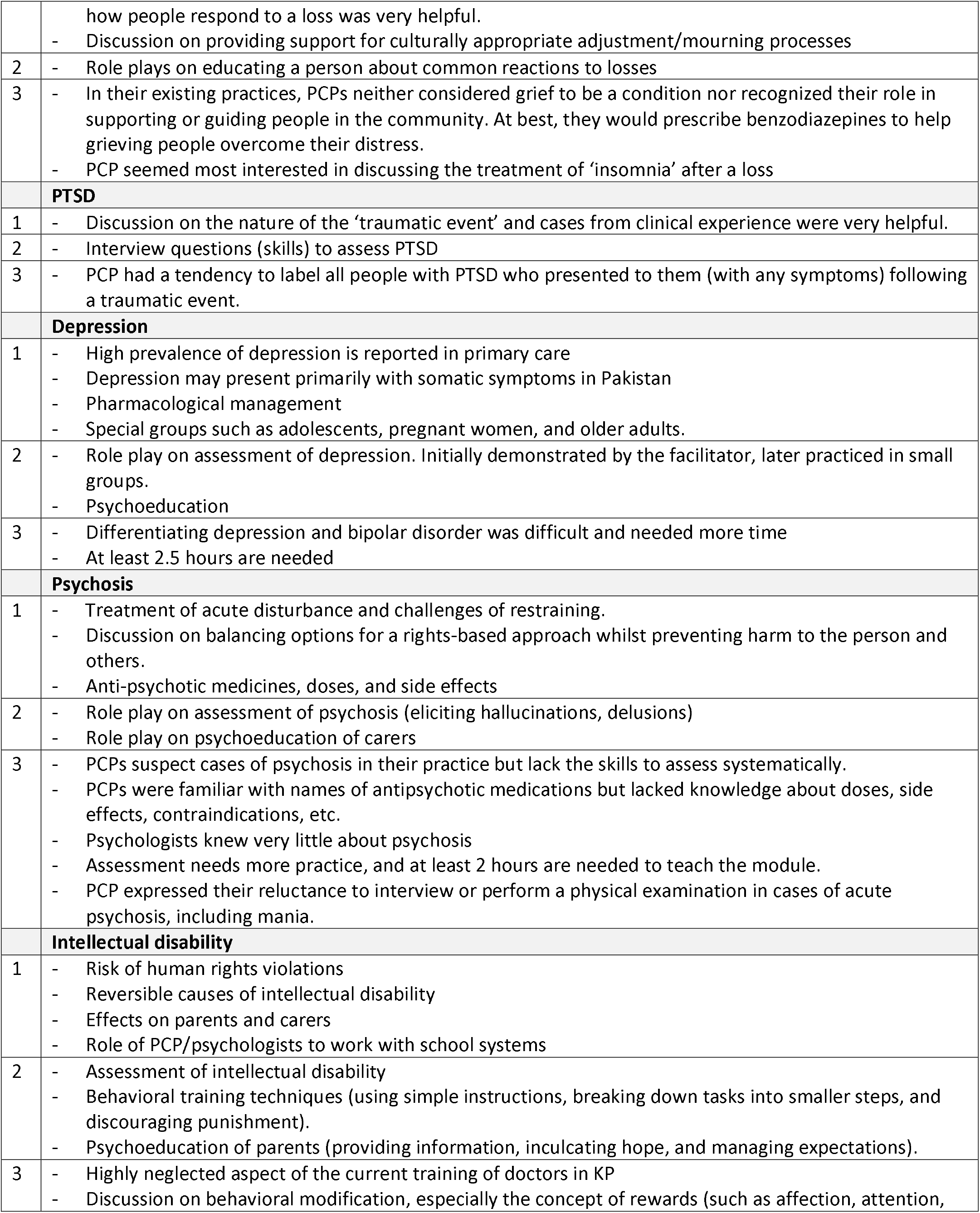
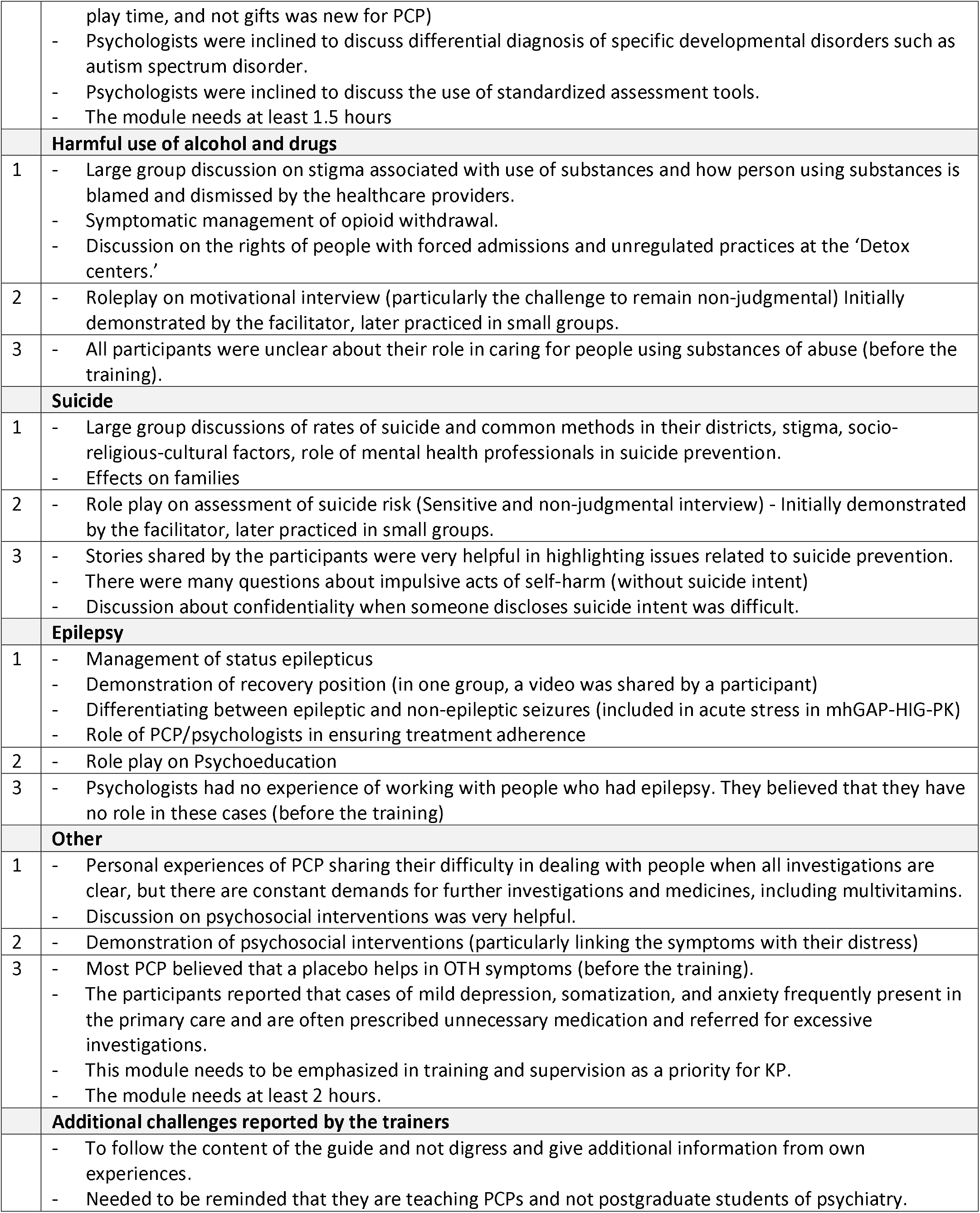
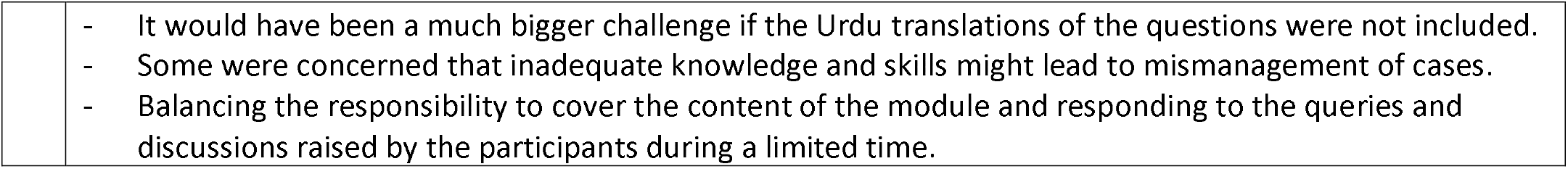
Content analysis of the feedback and reflections on individual modules.

1) Most beneficial aspect of training for enhancing knowledge identified by trainees.
2) Most valuable component of the training for strengthening skills reported by trainees.
3) Reflections by the trainers.

## Discussion

Although efforts to implement mhGAP-HIG in KP have been made in the last few years, these were isolated initiatives that lacked long-term planning and impact (20,21). The model of collaboration between the MoPD&SI (technical support), IMC (logistic and financial support), and the provincial health department (nominations of PCPs) is likely to help scale up the initiative effectively across the province.

Faregh et al. (2019) highlighted the challenge of identifying suitable trainers. Our experience suggests that incentivizing motivated mid-career specialists into a tailored career path could address this issue. Greater attention is also needed to select PCPs, as some primarily work as hospital administrators, others may lack interest in mental healthcare, and some close to retirement may not be interested in additional training. Furthermore, Pakistan’s under-resourced and challenging healthcare system has led many doctors to immigrate for better job opportunities. One study reported that a third of medical students aspire to move overseas after graduation owing to a lack of resources and ineffective management, impacting Pakistan’s socio-medical landscape (22,23). There is an imminent need to invest in younger, tech-savvy doctors who choose to work in their districts, interested in expanding their skills and more likely to have overcome stigmas associated with mental disorders.

The printed guide and mobile application proved to be valuable training tools, ensuring standardization across facilitators. The hard copies minimized reliance on technology and the internet, helped in better retention with personalized annotations, and enabled continuous learning outside formal training sessions. The guide’s Urdu interview questions and psychosocial techniques were particularly useful during role plays to build their confidence, improving skills to elicit symptoms, and offer psychosocial interventions. The trainers refrained from didactic teaching, and the trainees refrained from taking notes. The workshops became more interactive and hands-on as the trainees engaged proactively in clinical discussions and skills demonstrations. Participants recommended extending training duration to allow more case practice (9).

This was the first initiative in Pakistan involving both PCPs and CPs as trainers and trainees. Given growing number of trainings programs for CPs in Pakistan, they represent an underutilized mental health resource whose existing training gaps can be overcome through mhGAP training (24). The mixed group of participants facilitated a shift from a biomedical to a biopsychosocial approach as well.

During supervision, it became evident that referral pathways were influenced by the uneven distribution of specialist services across districts. The referral process was most effective when trainers worked in the tertiary hospitals, facilitating personal links between the PCPs and specialists. The absence of child and adolescent mental health specialists emerged as a critical gap in the referral system.

The training evaluation was also encouraging, where a substantial increase in knowledge was shown across all districts. The pre-test scores revealed notable knowledge gaps of common mental disorders, particularly in GPC, PTSD, SUI, SUB, and EPI. Significant Knowledge gaps in SUB and SUI were also found in Tunisia (25). There were three questions (T/F type) that were incorrectly attempted by two-thirds of the trainees in the pre-test, indicating widespread prevailing misconceptions. These included:

a. People with mental disorders cannot make decisions about their treatment/health.
b. People with drug dependence should be immediately admitted for detoxification.
c. Asking about thoughts of self-harm increases the risk of suicide.

Existing gaps in knowledge related to harmful use of substances have been documented in other humanitarian emergencies (26) According to United Nations Office on Drugs and Crime (2013), 10.7% of the population of KP “abused harder narcotics,” which was nearly double the national average at that time. Since then, there has been an alarming rise in the use of illicit substances, but health professionals are not equipped to respond with basic psychosocial interventions (27,28). Worryingly, this gap is steadily being filled by commercial and sometimes unscientific detoxification services.

The evaluation of our training is based on significant improvement in knowledge shown in the post-training testing, a trend that has been confirmed by other trainings (8,10,11,19). Logistic and budgetary constraints restricted our ability to explore knowledge retention across all districts at the 8 months. However, based on our analyses of six districts, we believe that knowledge retention at 8 months is likely to be consistent in the other districts, considering the demographic similarities, as well as immediate post-training results.

Our study demonstrates several strengths, including a collaborative approach, a diverse trainees’ group comprising PCPs and CPs, a highly interactive training experience, with a follow-up refresher training. Furthermore, the detailed daily reports prepared by trainers and summarized as ‘reflections’ proved valuable in identifying crucial areas within each module and highlighting the most effective methodologies. However, certain limitations were noted. Firstly, selection criteria for PCPs (based on clinical experience and interest) should have been specified. Secondly, trainers should have been formally engaged, preferably through financial or career incentives. Thirdly, our training pre- and post-evaluations could have been robust by using additional measures to assess skills, attitudes, and confidence. Lastly, the duration of training could have been extended as there was limited time for completing extensive content.

## Data Availability

All data produced in the present study are available upon reasonable request to the authors

## Future Directions

Mental health must be identified as a priority at the provincial level. A coordinating mechanism is needed between the health department, humanitarian agencies, and development partners. Currently, many projects are undertaken in silos with blurred and short-term outcomes. A clear direction needs to be set with focused objectives to optimize resource utilization.

An average district in KP has at least a hundred doctors working at the primary care level who are a huge potential resource for providing MHPSS services (29). Our work demonstrates a way forward to scale up capacity building initiative across the province. All efforts are needed to develop a multidisciplinary PHCWs workforce. For this reason, regular posts for CPs should be created at primary healthcare facilities.

Additionally, we recommend incorporating the guide into pre-service medical training, as its successful implementation in undergraduate and postgraduate programs for medical and allied specialties is well-documented (12,30).

## List of abbreviations

mhGAP-IG: Mental Health Gap Action Program,Intervention Guide;
mhGAP-HIG: Mental Health Gap Action Program,Humanitarian Intervention Guide; mhGAP-HIG-PK; Mental Health Gap Action Program,Humanitarian Intervention Guide Pakistan;
MHPSS: Mental Health and Psychosocial Support; MoPD&
SI: Ministry of Planning, Development & Special Initiatives;
PHCWs: Primary health care workers;
PCPs: Primary care physicians;
CPs: Clinical Psychologists;
IMC: International Medical Corps;
UNICEF: United Nations International Children’s Emergency Fund;
UNHCR: United Nations High Commissioner for Refugees;
LMS: Learning Management System;
PTSD: Post-Traumatic Stress Disorder;
FGDs: Focus Group Discussions;
ToT: Training of Trainers;
GBD: Global Burden of Disease;
YLDs: Years of healthy Life lost due to Disability;
ACU: Acute stress;
GRI: Grief;
ID: Intellectual Disability;
SUB: Harmful use of substances;
PSY: Psychosis;
ADHD: Attention Deficit Hyperactivity Disorder;
GPC: General Principles of Care;
COVID-19: Corona Virus Disease; LMICs;Low and Middle Income Countries;
OTH: Other significant mental health complaints;
MCQs: Multiple Choice Questions.

## Acknowledgments

We would like to thank Dr. Irshad Roghani, Director Public Health, Department of Health, Khyber Pakhtunkhwa and International Medical Corps, Pakistan for their support and collaboration.

We would also like to thank our team of trainers who contributed to the pilot testing without any incentives: Ibrahim Khan, Dr Izaz Jamal, Maria Ayaz, Dr M Muslim Khan, Shahzad Anwar, Dr Syed Tahir Hussain Shah.

## Ethics approval and consent to participate

Informed consent was obtained from members of focus group discussions. Participants were debriefed about the study. All participants voluntarily participated in the study. This study was conducted as part of the Mental Health and Psychosocial Support Project, approved by Ministry of Planning, Development and Special Initiatives, Government of Pakistan.

## Consent for publication

All authors consented to publication.

## Availability of data and materials

The data used in the current study will be available on request from the corresponding author.

## Author’s contributions

This study was conceptualized by AH, with AH also responsible for data curation and project administration. AN conducted the analysis, whereas AH and AN jointly devised the method and composed the original draft. The manuscript was edited and reviewed by AH.

