## Supplementary Table for "Implementing the mhGAP-HIG: The process and evaluation of training primary health care workers in Khyber Pakhtunkhwa, Pakistan" (from another version of this preprint: version 1, 2024-11-22)

*Figure 1: Quantitative analysis of the feedback*

|  | poor | fair | good | exc |
| --- | --- | --- | --- | --- |
| Overall Experience | 0.990099 | 0.990099 | 22.7723 | 75.2475 |
| Training Programme | 0 | 2.9703 | 22.7723 | 74.2574 |
| Facilitating skills of the trainers | 0 | 0.990099 | 9.90099 | 89.1089 |
| Training materials | 0 | 4.9505 | 31.6832 | 63.3663 |
| Methodology | 0 | 1.9802 | 21.7822 | 76.2376 |
| Opportunity to participate | 0 | 1.9802 | 36.6337 | 61.3861 |
| Peer learning | 0 | 1.9802 | 49.505 | 48.5149 |
| Logistics & planning | 2.9703 | 9.90099 | 50.495 | 36.6337 |
| Duration of training | 1.9802 | 12.8713 | 39.604 | 45.5446 |
